# Active Mutual Conjoint Estimation of Multiple Contrast Sensitivity Functions

**DOI:** 10.1101/2024.02.12.24302700

**Authors:** Dom CP Marticorena, Quinn Wai Wong, Jake Browning, Ken Wilbur, Pinakin Davey, Aaron R. Seitz, Jacob R. Gardner, Dennis L. Barbour

**Affiliations:** Department of Biomedical Engineering, Washington University, 1 Brookings Drive, St. Louis, MO 63130; Department of Computer Science and Engineering, Washington University, 1 Brookings Drive, St. Louis, MO 63130; College of Optometry, Western University of Health Sciences; Department of Psychology, Northeastern University; Department of Computer and Information Science, University of Pennsylvania

**Keywords:** Contrast sensitivity, psychophysics, machine learning, Bayesian modeling, active learning

## Abstract

Recent advances in nonparametric Contrast Sensitivity Function (CSF) estimation have yielded a new tradeoff between accuracy and efficiency not available to classical parametric estimators. An additional advantage of this new framework is the ability to independently tune multiple aspects of the estimator to seek further improvements. Machine Learning CSF (MLCSF) estimation with Gaussian processes allows for design optimization in the kernel, acquisition function and underlying task representation, to name a few. This paper describes a novel kernel for CSF estimation that is more flexible than a kernel based on strictly functional forms. Despite being more flexible, it can result in a more efficient estimator. Further, trial selection for data acquisition that is generalized beyond pure information gain can also improve estimator quality. Finally, introducing latent variable representations underlying general CSF shapes can enable simultaneous estimation of multiple CSFs, such as from different eyes, eccentricities or luminances. The conditions under which the new procedures perform better than previous nonparametric estimation procedures are presented and quantified.

**Precis:** Machine learning contrast sensitivity function estimation is improved by incorporation of additional information about the nature of the underlying and data from other eyes.

## Introduction

Visual contrast sensitivity functions (CSFs) represent a generalization of visual acuity testing capable of yielding insight into both peripheral and central visual processing. The main drawback of CSF testing for research and particularly for clinical applications is long acquisition times. A standard compromise to achieve informative CSF estimates in practical amounts of time is to make strong assumptions about the nature of the CSF, i.e., its parametric form. Optimal data acquisition procedures under these assumptions result in efficient and practical testing procedures (Gu et al., 2016; Lesmes et al., 2010; Wang et al., 2016). Estimation accuracy for unusual CSF phenotypes suffers in the process, however (Chung & Legge, 2016; Rohaly & Owsley, 1993; Tahir et al., 2009; Woods et al., 1996).

Conventional wisdom indicates that relaxing model assumptions to allow more flexibility would enable higher accuracy at the expense of lower efficiency. We showed previously that a flexible, nonparametric Bayesian estimator exhibits a compelling accuracy/efficiency tradeoff compared to classical CSF estimation strategies (Marticorena et al., 2024). Perhaps more importantly, this machine learning CSF (MLCSF) estimator has the added benefit of many options for further refining this and other tradeoffs in order to improve its performance.

The first-generation MLCSF estimator, while reliable, had particular drawbacks. While it showed an improved ability to estimate unusual CSF shapes compared to a standard parametric estimator, it was challenged to fully capture behavior at the highest spatial frequencies at the same time. Additionally, the stimuli selected for delivery by strictly optimization were often not well-distributed along spatial frequency, often becoming concentrated at lower spatial frequencies. This lower density sampling at higher spatial frequencies appeared to increase the variance of final CSF estimates across repeated runs. Finally, the efficiency of MLCSF was overly sensitive to low-probability labels (i.e., a miss or a successful guess) at early stages of learning, as several nearby stimuli were typically needed to counter a mislabeled data point.

Thus, it became apparent that some combination of improved stimulus selection (i.e., data sampling) and model regularization might combine to improve both accuracy and efficiency of a second-generation MLCSF estimator.

In this article we examine the ability to improve active MLCSF estimation upon relaxing two standard constraints: 1) strictly monotonic likelihood in contrast and 2) new stimulus selection using strictly maximum information gain. As a third manipulation, we explore the potential benefits of estimating two CSFs simultaneously, as an informed regularization method. This conjoint estimation procedure has been shown to accelerate final estimate convergence in psychometric fields even between dissimilar phenotypes (Barbour et al., 2018).

## Modeling Framework

### Background

As in previous work, the conceptual starting point for a machine learning CSF estimation procedure is a probabilistic classifier that separates two regions in feature space designated “success” and “failure” (Marticorena et al., 2024; Song et al., 2015). Success for a contrast detection task is defined as a button press immediately following the delivery of a Gabor stimulus of a given spatial frequency and contrast. Failure is defined as exiting the response window following stimulus delivery without a button press. A probabilistic classifier is preferred over a true classifier because the boundary between success and failure in the former case is gradual rather than discrete, consistent with signal detection theory.

Further, a probabilistic classifier is a natural framework to estimate a full psychometric field, which maps success probability for all combinations of independent variables (i.e., features), resulting in a generative model of item-level predictions (Song et al., 2018). In the present case, this is the Contrast Response Function (CRF), which takes the form of a probability surface defined over spatial frequency and visual contrast. If all non-psychometric independent variables are held constant (i.e., spatial frequency for the CRF), a psychometric field reduces to the familiar psychometric curve, in this case reflecting the probability of detecting a visual stimulus at any contrast value. A predetermined probability of detection (e.g., 50%) is the single point on the psychometric curve defining the threshold. Evaluating this threshold over all spatial frequencies contributing to the CRF yields the CSF.

The Gaussian process (GP) used to represent the Machine Learning CRF (MLCRF) is an inherently flexible, nonparametric modeling framework. It offers many potential options for customization that have the potential to improve estimation. Of particular interest is the ability to finely control the inductive bias (i.e., the modeling assumptions and their confidence) through several different means. Incorporation of a Bayesian informative prior, for example, has the potential to speed estimation, although it was not found to do so systematically under the MLCSF estimation conditions tested previously (Marticorena et al., 2024). In this study we evaluated the contributions of the 1) GP kernel, 2) acquisition function and 3) multitask GP configuration toward improving MLCRF and MLCSF estimation accuracy and efficiency.

### Gaussian process classification

A full development of the following can be found in (Marticorena et al., 2024). Briefly, define *f*(**X**) to be a latent function over a continuous multidimensional feature space **X** ∈ *χ*. A Gaussian process (GP) is a convenient way to encode prior knowledge about the latent function:

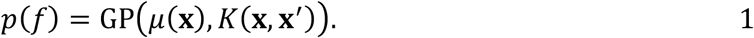

This knowledge, encapsulated within the mean function μ and the covariance function or kernel *K*, can be updated following new data collection according to Bayesian principles.

The GP is trained over a finite sample of independent features **X** = {**x**_**1**_, **x**_**2**_,…, **x**_7_}. In binary classification tasks the dependent variable can take on one of two values indicating failure or success: *y*_*i*_ ∈ {0,1}. The latent function models this response-generating function, resulting in the MLCRF. Dividing the input domain of the psychometric field into subdomains of “task success” and “task failure” recasts the traditional signal detection regression problem in terms of classification, allowing advances in powerful machine learning classification algorithms to be exploited.

### Kernel tuning

In the first-generation MLCRF estimator, the probability of success *p*(*y* = 1|*f*) was modeled by a sigmoidal link function *ψ*(**x**) consistent with signal detection theory. This function is distributed according to a Bernoulli likelihood:

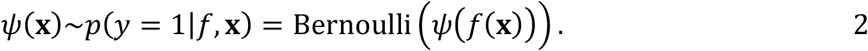

Therefore, a linear kernel observed through the link function results in an inhomogeneous Bernoulli likelihood monotonic in contrast. The form of the link function accommodates mislabeled responses:

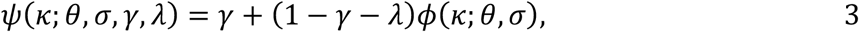

where *ϕ*(·) is a cumulative distribution parameterized by threshold *θ* and spread *σ, γ* is a guess rate, and *λ* is a lapse rate (Wichmann & Hill, 2001). The independent variable *κ* represents contrast.

A linear kernel operating on contrast alone reflects the assumption of monotonicity in contrast. To accommodate an assumption of local smoothness in spatial frequency, a squared exponential kernel was originally used, operating on spatial frequency alone:

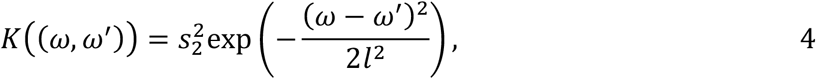

where *s*_2_ represents a scaling factor and *l* represents a length constant, which governs the smoothness of the function. The independent variable *ω* represents spatial frequency and *ω* ′ indicates any other spatial frequency to compute the covariance against.

The overall kernel for the first-generation MLCRF was a simple linear combination of the constituent kernels (Marticorena et al., 2024):

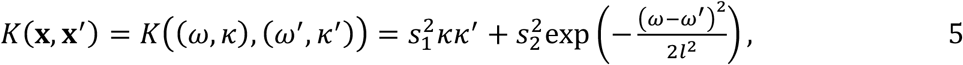

where **X** = (*k, ω*). Performance of the original MLCSF estimator indicated that while overall accuracy was good, threshold values at very high spatial frequencies were less accurate than average for some phenotypes. In particular, CSFs with very steep falloffs at high spatial frequencies required shorter squared exponential length constants to fit accurately, leading to inappropriate rippling at lower spatial frequencies, thus lowering overall accuracy. Conversely, if the lower spatial frequencies were fit well, accuracy at higher spatial frequencies may suffer due to longer squared exponential length constants.

An alternate kernel design strategy involves relaxing the contrast constraint in order to obtain more flexibility for spatial frequency. The intent is to achieve good fits at both high and low spatial frequencies. This functionality is achieved by allowing contrast to have its own smoothness term in addition to the linear term of before. This second-generation kernel can be given as follows:

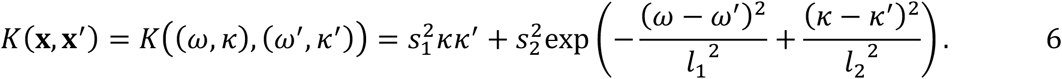

### Acquisition function tuning

Bayes’ rule is applied to compute updated posteriors upon observation of data {**y, X**}. Because of the Bernoulli likelihood, the posteriors cannot be computed in closed form. They are estimated in this case via variational inference (Hensman et al., 2015; Titsias, 2009). Variational inference finds the best approximation of the true posterior distribution from a family of simpler distributions by minimizing the Kullback-Liebler divergence between the approximate and true posteriors (Gardner et al., 2018).

We can efficiently estimate the posterior distribution of the GP model, which when trained with all existing data can compute model updates for any new sample **x**^∗^ ∈ **X**^∗^ defined over spatial frequency and visual contrast. Therefore, the new sample **X**^∗^ that, upon observation, maximizes some utility function *U*(**x**^∗^) is optimal under that function:

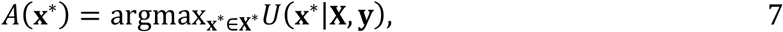

where *A*(·) represents the acquisition function and *U*(·) is a utility function reflecting model quality. Previous implementation of the first-generation acquisition function prioritized uncertainty sampling by defining the utility function as the differential entropy calculated using the predictive mean *μ* and variance *σ*^2^ of the model observed through the likelihood (Marticorena et al., 2024):

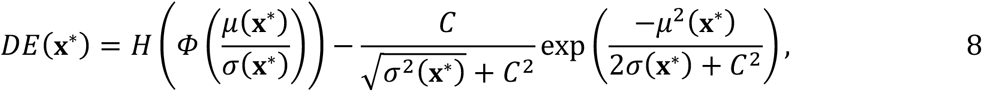

where *H* is the binary entropy function *H*(*x*) = −*x*log_2_(*x*) − (1 − *x*)log_2_(1 − *x*), Φ is the CDF of a standard normal, and *C* is a normalizing factor 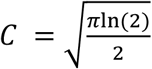, which affords the approximation of the second term in closed form (Houlsby et al., 2011). This acquisition function finds the next best sample point **X**^∗^ that maximizes the differential entropy, which is a proxy for information gain.

The gradients of differential entropy for the first-generation MLCRF estimator were very similar over contrast and very shallow over spatial frequency. Imposing a hard nonlinearity under these conditions can lead to degenerate conditions in which the updated model changes little, leading to multiple samples in close proximity. We hypothesized that implementing a sampling density penalty would provide a better balance between exploration and exploitation than the original acquisition function allowed.

The second-generation acquisition function, rather than taking the maximum of *U*(**x**^∗^|**X, y**), prioritizes a subset, **X**_*hi*(*n*)_, which are the points within the highest *n*%. Next, it applies nearest neighbors to identify the point **x**^∗^, within **X**_*hi*(*n*)_ that is furthest from any point in the existing sample set **X**. The new sampling density penalty in the acquisition function allows for more effective exploration initially, preventing proximity oversampling. As data acquisition continues, the new method enhances exploitation, ensuring more uniform sampling across spatial frequencies.

### Conjoint estimation

The discussion above corresponds to a single GP defined over a feature space consisting of the independent variables of the psychometric field, resulting in a single MLCRF estimate. To estimate multiple GP models simultaneously, we extend our modeling framework and acquisition function over an augmented feature space for two eyes or experimental conditions. This can be accomplished in multiple ways, including kernel extension to learn different pairwise correlations for observations made in combinations of the individual feature spaces (Barbour et al., 2018). In that case, conjoining hyperparameter(s) are learned alongside other model hyperparameters. When data are acquired under active learning conditions for two phenotypes, conjoint estimation can halve the data requirements for accurate estimation, even when the phenotypes differ (Heisey et al., 2018).

For the current study we implement conjoint estimation of two MLCRFs as a MultiTask GP using the linear model of coregionalization (Goulard & Voltz, 1992). This approach enables the modeling of multiple related tasks in a cohesive structure, where each task, *f*_*task*_(*x*), is modeled as an output dimension of a linear combination of a set of latent functions, *g*(⋅) = [*g*^(1)^(⋅), …,*g*^(*Q*)^(⋅)].

Each latent function is represented by an independent GP. The relationship is given as:

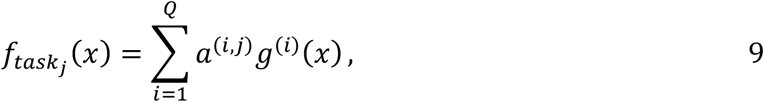

where *a*^(*i*)^ are learnable parameters. This configuration facilitates the simultaneous learning of distinct sets of variational parameters and hyperparameters for each latent function.

Because each MLCRF must be characterized by at least one latent function, the number of latent functions (*Q*) must be equal to or greater than the number of MLCRFs. The second-generation model sets the number of latent functions to be equal to the number of MLCRFs, which is 2 for the current experiments. Note that this implementation does not reduce the model to 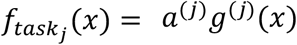.

## Methods

### Simulations

In experiment 1, ground truth CSF models for four canonical phenotypes were constructed from idealized threshold curves (Kalloniatis & Luu, 1995; Marticorena et al., 2024). In experiment 2, ground truth CSF models were drawn from 8 experimental conditions for 9 individuals (Jigo & Carrasco, 2020).

In all cases, threshold curves as a function of spatial frequency were simulated by cubic spline interpolation and quadratic extrapolation. The resulting threshold curves were used to create generative ground truth CRF models representing a visual pattern-detection task as a function of spatial frequency and visual contrast. At each spatial frequency a one-dimensional sigmoidal psychometric curve was constructed using the four-parameter model in equation 3. One difference for the generative model, however, is that the sigmoid is given by the standard logistic function

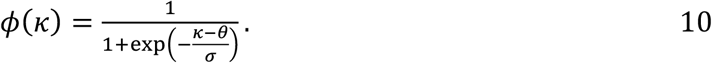

At any contrast value *κ*, binary observations (i.e., detected or not detected) were generated by sampling from a Bernoulli distribution with success probability given by *ψ*(*κ*) = *p*(*y* = 1|*κ*).

### Psychometric field for contrast response

The psychometric field defining the item-level generative CRF model over the domains of spatial frequency and contrast is given by *ψ*(*ω, κ*), while the underlying continuous latent function is defined by *f*(*ω, κ*). The estimation procedure consists of acquiring the best data to learn model hyperparameters, which for the GP are given by the closed-form representations of the mean function and covariance function or kernel (Rasmussen & Williams, 2006).

In all models for the current study, the GP mean function is given as a constant:

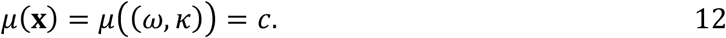

This simple formulation purposefully imbues the kernel with most of the learning.

First-generation estimators enforce strict linearity in contrast and smoothness in spatial frequency, as indicated in equation 5. For second-generation estimators the kernel encourages linearity and smoothness in contrast and smoothness only in spatial frequency, as indicated in equation 6.

For first-generation estimators the acquisition function maximizes information gain, as seen in equation 7. Second-generation estimators generalize the acquisition function to include a sampling density consideration:

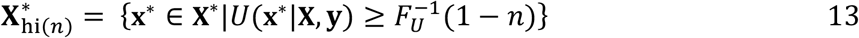

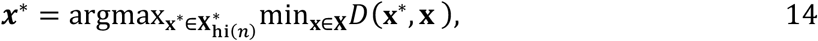

where 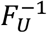 is the inverse cumulative distribution function, *n* is a percentile constant, and *D*(**x**^∗^, **x**) is the distance between each candidate point **x**^∗^ and its corresponding nearest point in the previously collected data **x**. Thus, the acquisition function selects the point **x**^∗^ that, among points in 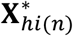, has the greatest minimum distance to any given existing sample point.

### Implementation

All simulation, machine learning, and evaluation software was written in Python using major libraries Python (*Python 3.10.9*, n.d.), PyTorch (*PyTorch 1.13.1*, n.d.) and GPyTorch (*GPyTorch 1.8.1*, n.d.). Project information, including code and data necessary to replicate these experiments, can be found at https://osf.io/7ujga/.

Two experiments were conducted in order to test the new estimator configuration under different conditions. For experiment 1, four ground truth generative models were created from four canonical CSF phenotypes as in (Marticorena et al., 2024). Fifteen logarithmically spaced values were used along each of 6 octaves of spatial frequency (*ω* ∈ [1, 64] cycles per degree) and 30 values for each of 3 octaves of contrast (*κ* ∈ [0.001, 1]), resulting in a 91×91 grid of logarithmically spaced values. This resolution is arbitrary and can be modified as needed. Cubic spline interpolation and quadratic extrapolation were used to determine continuous CSF values at every spatial frequency. This CSF curve was combined with a plausible psychometric spread value of 0.08 everywhere to create a generative CRF (Zhao et al., 2021).

For each phenotype, data acquisition always began with a fixed primer sequence of two points: one at the origin of the grid (i.e., the lowest spatial frequency and lowest contrast on the grid), and an additional point at the first octave division of *κ* and third octave division of *ω*. Active learning then commenced as determined by the acquisition function. The GP estimator models determined the next features to sample, and the generative models provided probabilistically determined labels of success or failure. Data acquisition for each eye or pair of eyes terminated following 100 or 200 total samples, depending on the protocol.

Conjoint estimation take on a particular formulation for these experiments referred to as active mutual alternating conjoint estimation. In this case, odd stimuli are delivered to one eye and even stimuli to the other eye. After each stimulus delivery, posteriors for both eyes are updated and the next stimulus for the other eye determined strictly from the analysis of its utility function.

The Cataract and Normal phenotypes were selected for an in-depth evaluation of the 4 experimental conditions, as these phenotypes occupy opposite extremes of phenotype space. The *ϕ* = 0.5 contour of the CRF predictive posterior mean became the CSF estimate. The root mean square error (RMSE) between the ground truth CSF and the estimated CSF in log contrast units was quantified at all spatial frequencies. The estimated CSF was discretized to the nearest contrast grid value for this calculation. Each phenotype pair was evaluated separately for 25 repetitions with different random seeds and the overall mean behavior summarized.

Experiment 2 made use of generative ground truth models taken from a cohort of 9 individuals performing a large number of contrast sensitivity detections at a variety of eccentricities and attentional conditions. These highly oversampled, and thus highly confident, CSF phenotypes determined from the original threshold estimation procedure are depicted in **Figure 1** (Jigo & Carrasco, 2020). Another 91×91 feature domain grid of logarithmically spaced values similar to Experiment 1 was used, except the spatial frequencies were estimated over the domain of *ω* ∈ [0.5, 32] cycles per degree. Because only discrete spatial frequencies were evaluated in the original experiments up to 11 cycles per degree, quadratic extrapolation was used to represent each CSF curve at higher spatial frequencies. Other experimental conditions were identical to experiment 1.

**Figure 1:**
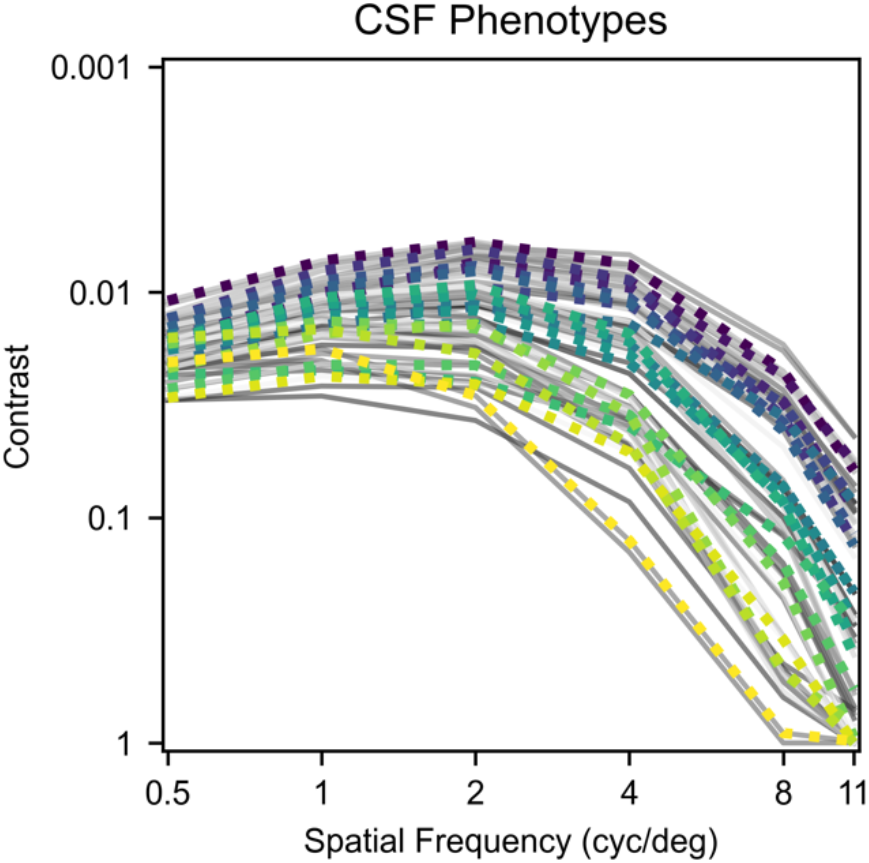
The 72 different phenotypes evaluated from a population of 9 individuals under different stimulus conditions considered for generative models for experiment 2 (Jigo & Carrasco, 2020). Thicker dotted lines indicate 20 representative CSF curves sampled for further analysis. These 20 curves, equally spaced in rank, form ventiles.

There are 2628 unique pairings for the 72 CSF phenotypes from the human data. Rather than evaluate all of them, averaged Pearson correlation coefficients were computed between each original CSF and the remaining 71. These averages were divided into 20 equally spaced ventiles. From each ventile, the curve with the median correlation was selected, yielding 20 representative curves encompassing the dataset’s diversity. These curves are highlighted in color in Figure 1. Estimation performance was averaged across all 210 unique pairings of these select phenotypes.

For all estimator models, the mean function of the GP was initialized with *c* = 0. Zero on the latent function maps to a probability of success of 0.5, implying that without any data, the estimator assumes maximum uncertainty about the shape of the CRF. The kernel was initialized such that all hyperparameters were initially assigned a value of 1. The intention of this prior was to allow the sampled data to speak for itself in order to deliver a final estimate with few assumptions. Previous testing has not yet revealed a superior prior for CSF estimation (Marticorena et al., 2024). In every condition, a set of phantom shaping data points were added to assist estimator convergence. These values indexed detection failures at stimulus configurations well beyond any reported human contrast sensitivity. At each octave of spatial frequency, a phantom failure at a contrast of 0.0005 was added. Another phantom failure was added at a spatial frequency/contrast combination of (128, 1).

### Evaluation

The MLCRF estimator generates a predictive posterior mean as a function of spatial frequency and contrast, which is equivalent to a maximum *a posteriori* estimate. The 0.5 probability counter as a function of spatial frequency was extracted to represent the MLCSF estimate. For each spatial frequency, the deviation of MLCSF from ground truth was quantified by RMSE over contrast. This summary of model quality was updated after each new sample point, allowing comparisons between experimental conditions in RMSE evolution plots.

## Results

### Experiment 1

A comparison of disjoint and conjoint estimation in model configuration 1 for twin Normal phenotypes can be seen in **Figure 2**. Following 50 actively acquired stimulus presentations per paired phenotype (100 per pair), credible CSF estimates are provided for all 4 individual phenotypes. For disjoint estimation shown in the top row, the models are essentially independent repeats of the same estimation procedure. The two estimated CSF curves are indicative of the variety of forms that first-generation MLCSF takes, including rippling at lower spatial frequencies. Conjoint estimation with the same model configuration in the bottom row, however, results in visibly higher accuracy at 50 samples. Closer inspection reveals that more of the conjointly-acquired stimuli are closely approximated to the ultimate CSF curve estimate, implying a more rapid convergence toward the final estimate than disjoint estimation produces.

**Figure 2:**
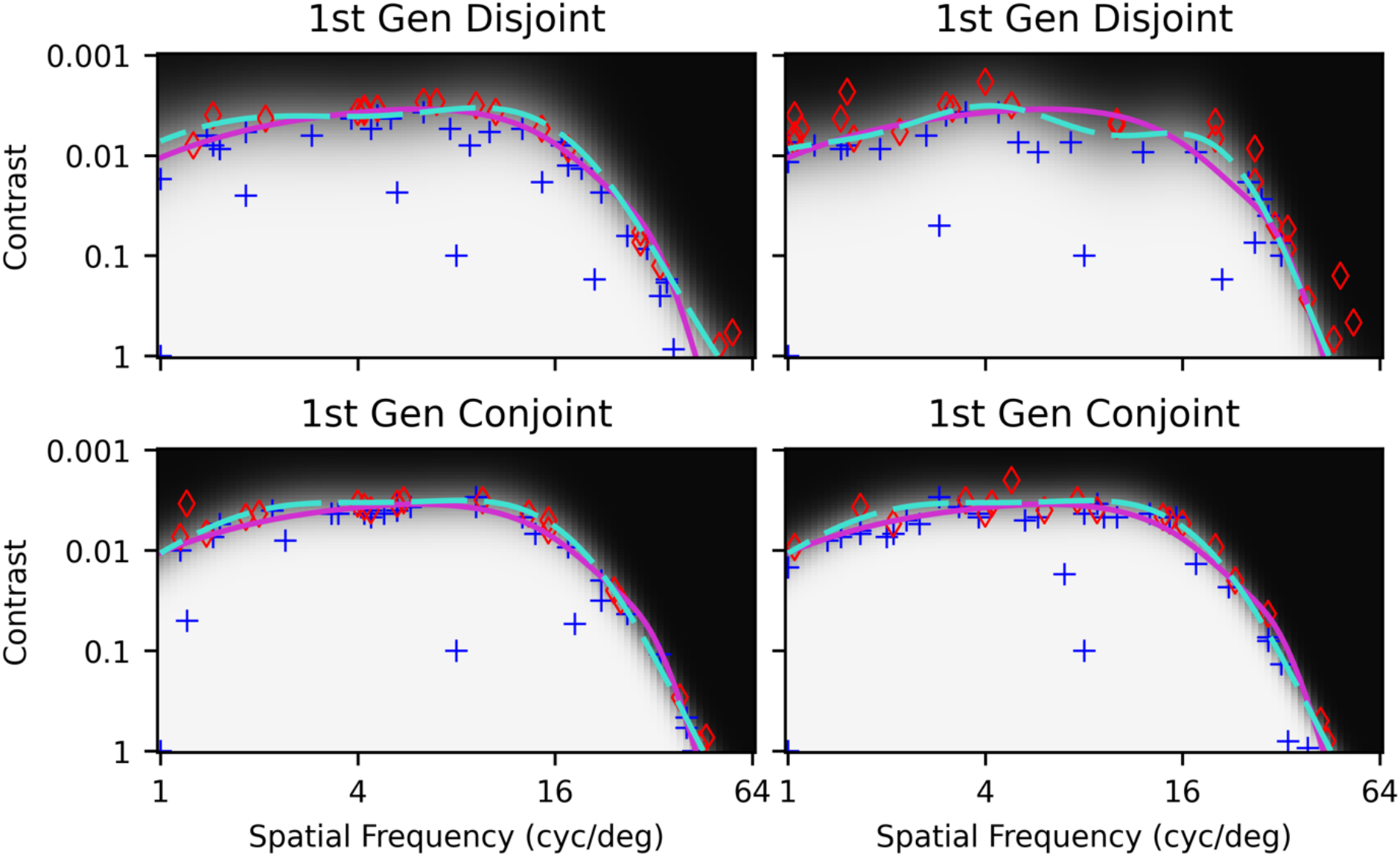
First-generation disjoint (top) and conjoint (bottom) MLCRFs estimated with 50 samples of active learning per phenotype (100 total) for a (Normal, Normal) canonical phenotype pair. Simulated behavioral responses (blue plus signs = success, red diamonds = failure), ground truth Normal CSF (solid magenta curve), learned MLCSF (dashed cyan curve) and predictive posterior mean values (grayscale) are also plotted. Top RMSE values: 0.101, 0.112. Bottom RMSE values: 0.0920, 0.0908.

One potentially important difference between CSFs estimated disjointly versus conjointly for the first-generation kernel is that the latter make fewer systematic errors in threshold determination at particular spatial frequencies. This result comes about because the conjoint estimates are more regularized and thus smoother on average. Disjoint estimates often either end up rippling at low spatial frequencies with steeper estimates at higher spatial frequencies, or have smoother estimates at lower spatial frequencies with shallower estimates at higher spatial frequencies.

Conjoint estimates, on the other hand, more frequently strike a middle ground between avoiding low-frequency rippling in addition to yielding acceptable slopes at higher spatial frequencies.

Also noticeable is the occasional clustering of some stimuli quite close together, often at low spatial frequencies. The first-generation acquisition function is simply an information optimization, with no explicit component promoting a wide distribution of stimulus samples. Strict optimization of information gain can lead to such clustering under conditions in which the utility function changes little between samples.

These two observations were made following the initial comparison between disjoint and conjoint MLCSF estimation, leading to the hypothesis that compensating for them directly might improve overall estimation accuracy and/or efficiency. For this reason, a new estimator was designed that relaxed constraints on the overall CSF shape while placing greater constraints on the distribution of stimulus samples to be delivered. Specifically, stimulus samples were selected such that similarity between any two stimuli was reduced. The result was the second-generation estimator MLCSF estimator.

Figure 3 reveals the effects of these manipulations, once again for 50 samples per phenotype (100 per pair). All 4 MLCSF curves closely approximate the underlying ground truth models. Even though the second-generation MLCSF is free to curve back on itself at higher spatial frequencies with the new kernel, that occurrence is generally prevented by the particular selection of stimulus samples delivered. The wider sample distribution and lack of any clustering is apparent and may be contributing to improved estimation accuracy in this case. Judging from the few stimuli far from threshold, selection of stimuli close to threshold happens at least as quickly as the first-generation example and probably faster.

**Figure 3:**
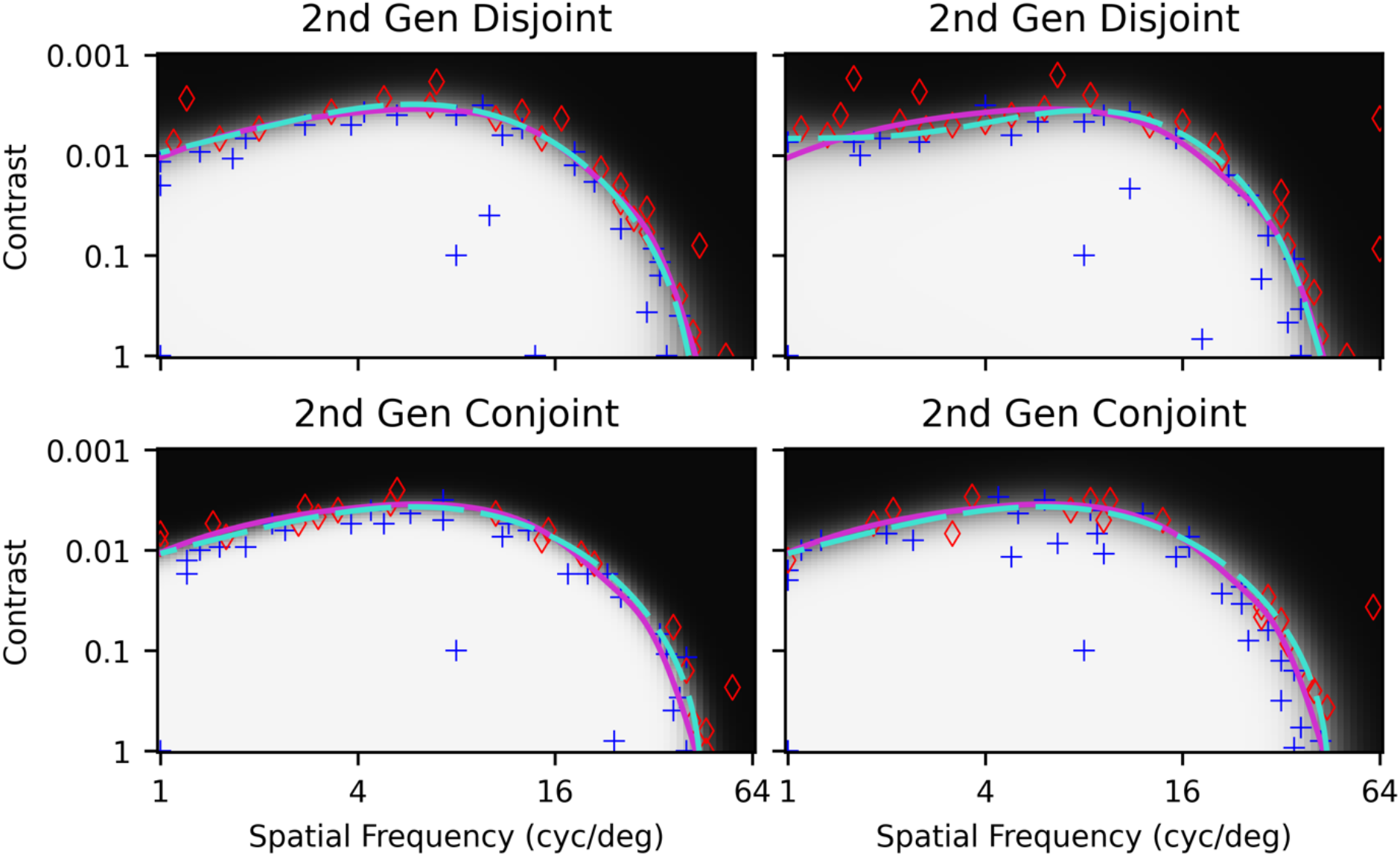
Second-generation disjoint (top) and conjoint (bottom) MLCRFs estimated with 50 samples of active learning per phenotype (100 total) for a (Normal, Normal) canonical phenotype pair. Plotting conventions are as in Figure 2. Top RMSE values: 0.0458, 0.0862. Bottom RMSE values: 0.0877, 0.0877.

This estimator property has a synergistic effect on estimation accuracy. The guess and false positive rates for this detection experiment were 4%. When a mislabeled data point emerges, active learning selects several more similar stimuli in order to ensure the label is an error and not a surprise participant response. By converging toward threshold extremely rapidly—even without an informative Bayesian prior to shape the model early on—the average informativeness of each stimulus toward threshold determination increases, thus improving efficiency. The more distributed (i.e., less clumped) samples from the second-generation acquisition function appear to aid this rapid convergence.

To gain a better understanding of the utility of the second-generation estimator configuration, it was compared directly against the first-generation configuration for both disjoint and conjoint estimation, as well as a classical CSF estimator for comparison (Canare et al., 2019). Average MLCSF estimation accuracies as a function of the amount of data actively collected for three different phenotype pairings are depicted as RMSE evolution plots in **Figure 4**. In the (Normal, Normal) pairing (top row), active alternating mutual conjoint estimation tends to achieve higher accuracy at lower sample counts than disjoint estimation. At higher sample counts this disparity mostly disappears, indicating that all estimators are converging toward consistent estimates. The variation in conjoint estimates is systematically lower than in disjoint estimates, resulting in higher precision than the disjoint case. The poor precision of the first-generation disjoint case resulted from a single example that failed to converge.

**Figure 4:**
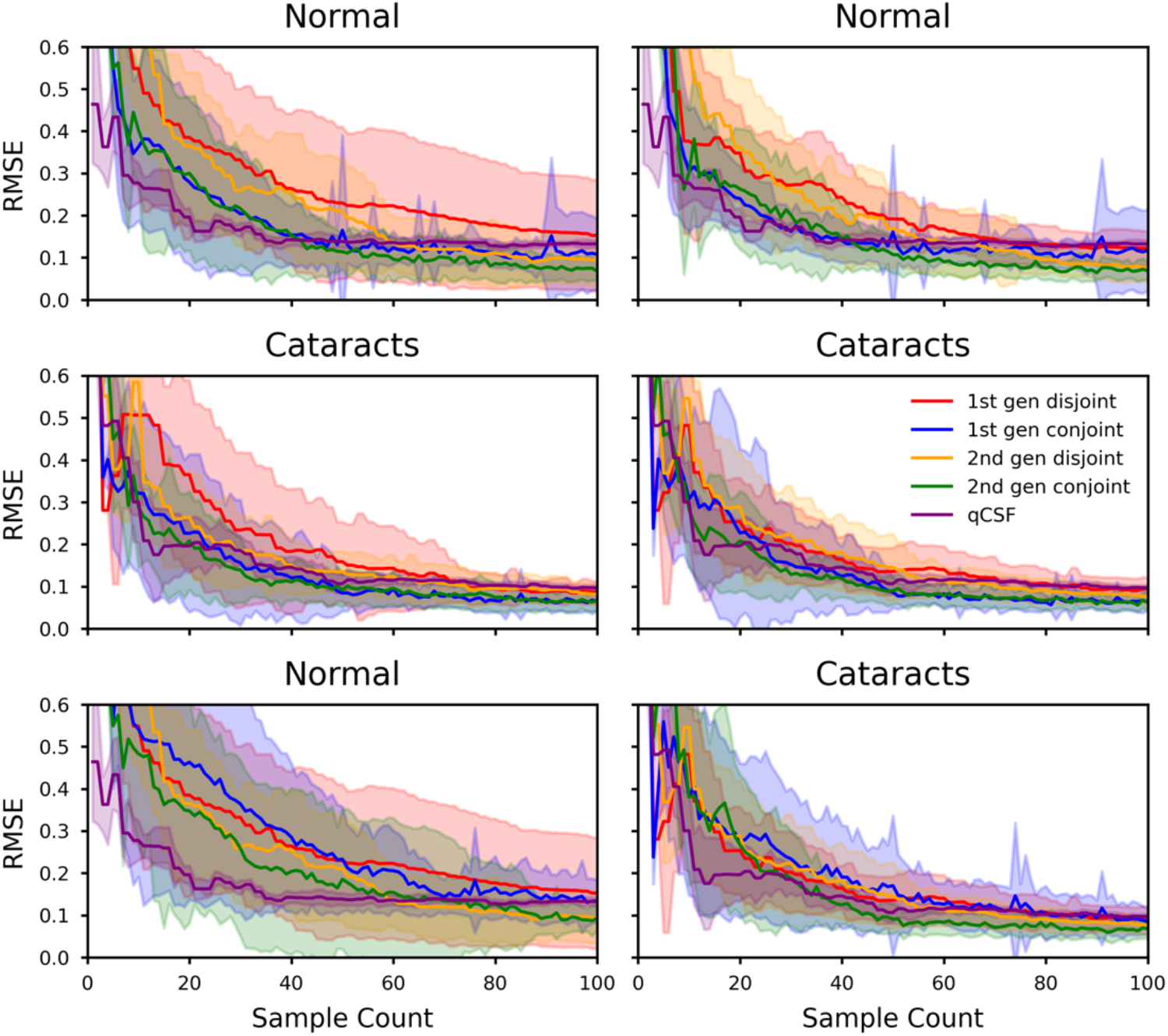
Mean ± standard deviation RMSE values in log contrast units for pairs of phenotypes averaged from 25 repeat experiments using up to 50 samples per phenotype (100 per pair). Curves reflect first-generation disjoint MLCRF (green), first-generation conjoint MLCRF (purple), second-generation disjoint MLCRF (blue), second-generation conjoint MLCRF (red), and for comparison, an implementation of quick CSF (purple). Shaded polygons reflect estimate precision. Data were actively sampled across both members of each pair, for (Normal, Normal) (top), (Cataracts, Cataracts) (middle), and (Normal, Cataracts) pairs.

To compare disjoint and conjoint estimators in Figure 4, models for both phenotypes were updated each time a new sample was acquired. Because in the disjoint case this new sample had no way of changing the other phenotype, the model for that phenotype was copied and repeated one sample forward, rather than being re-estimated. As a result, the disjoint RMSE evolution curves take on a staircase appearance. This same approach was taken for the quick CSF curve. The total amount of data reflected for the complete model for each phenotype in these plots is 50 samples. The total amount of data for both models is 100 samples.

A substantially similar result can be seen for the (Cataract, Cataract) condition in the middle row of Figure 4. Across model versions, conjoint estimation systematically converges faster compared to disjoint, while at the same time achieving obviously improved final estimates, irrespective of model version. The main difference from (Normal, Normal) is that first-generation conjoint MLCSF performs as well as second-generation conjoint MLCSF for the (Cataract, Cataract) condition. Regardless of the model configuration, conjoint estimation is showing efficiency and consistency performance at least as good as disjoint estimation and, in some conditions, better.

The previous two conditions involved learning models for two identical phenotypes. This condition reflects the inspiration for the original conjoint estimator design because many experimental conditions emerge in which two models are expected to be quite similar. A major surprise in previous work, however, was finding that even when the two phenotypes differed substantially, performance gains still occurred (Barbour et al., 2018; Heisey et al., 2018).

Presumably enough similarity between even highly dissimilar phenotypes exists for an estimator learning those correlations (rather than assuming independence) to leverage them for improving estimation accuracy with fewer data.

An interesting result is observed with the (Normal, Cataract) pairing in the bottom row of Figure 4. Within model versions, the conjoint estimation advantage is no longer as obvious as with the previous conditions. The only obvious trend across estimation of the disparate phenotypes is that second-generation conjoint MLCSF, in particular, converged more rapidly and with the best final estimates.

Note that first-generation conjoint MLCSF estimation occasionally updates to a less accurate model, as can be seen in the occasional sharp peaks in the top and bottom rows of Figures 4. This type of momentary model unlearning does not tend to occur in the disjoint condition, nor in the second-generation conjoint MLCSF, leading to its designation as “negative inflexible conjoint interference.” Fortunately, the estimation process always recovers from these problematic model updates and goes on to deliver an accurate MLCSF estimate. Furthermore, the second-generation conjoint estimator appears not to be susceptible to negative conjoint interference, likely due to its increased flexibility.

All previous analyses have been computed as RMSE vertically along contrast, averaged across all spatial frequencies. Average MLCSF estimation accuracy evaluated as a function of spatial frequency of data actively collected for a (Normal, Normal) phenotype pairing are depicted in **Figure 5**. The first-generation kernel, particularly with disjoint estimation, is susceptible to systematically greater, worsening error at higher spatial frequencies. This high-spatial-frequency error can be seen at values as low as around 8 cycles/degree, with maximal error reliably at the *κ* = 1 intercept (i.e., the point of maximum contrast on the CSF). While conjoint estimation reduces these effects for the first-generation model, they are still strong and obvious. While the second-generation model also exhibits worsening error at higher spatial frequencies, it is substantially lower than the first-generation kernel, and generally low below 32 cycles/degree.

**Figure 5:**
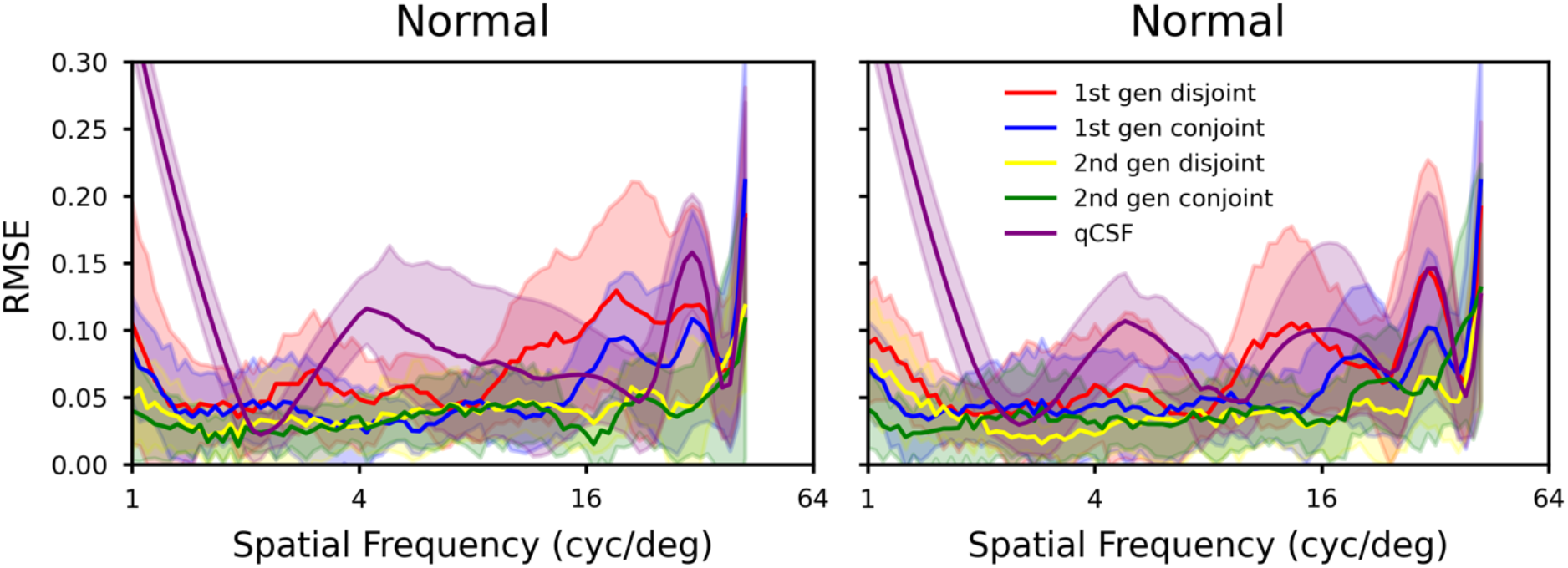
Mean ± standard deviation RMSE errors in cycles per degree of a (Normal, Normal) phenotype pair, averaged from 10 repeat experiments using 100 samples per phenotype. Data sources were as in Figure 4.

### Experiment 2

Experiment 1 revealed that the second-generation MLCSF with its more flexible nature did not systematically worsen estimation performance and appeared to combine favorably with conjoint estimation procedures to produce consistently good results. Therefore, the second-generation configuration was selected for further experimentation. In order to obtain more realistic performance estimates, oversampled human data were reconstituted into simulated individuals, and CSFs were estimated for 210 distinct pairs, chosen from 20 phenotypes distributed across correlation quantiles within the population (Figure1).

The result of these experiments can be seen in **Figure 6**. The larger number of examples in this case leads to smoother and more obvious trends. The conjoint estimator is statistically significantly more accurate than the disjoint estimator at 100 overall samples (*p*_1000000_ < 1.0×10^−6^, *n* = 420, permutation test). That difference disappears by 200 overall samples, however (p_1000000_ = 0.593, *n* = 420, permutation test). Therefore, conjoint estimation under these experimental conditions requires significantly fewer data to achieve accurate final fits. Furthermore, the variance across pairs is also statistically significantly smaller in the conjoint condition with 100 overall samples (*p*_1000000_ < 1.0×10^−6^, *n* = 420, permutation test), indicating greater consistency.

**Figure 6:**
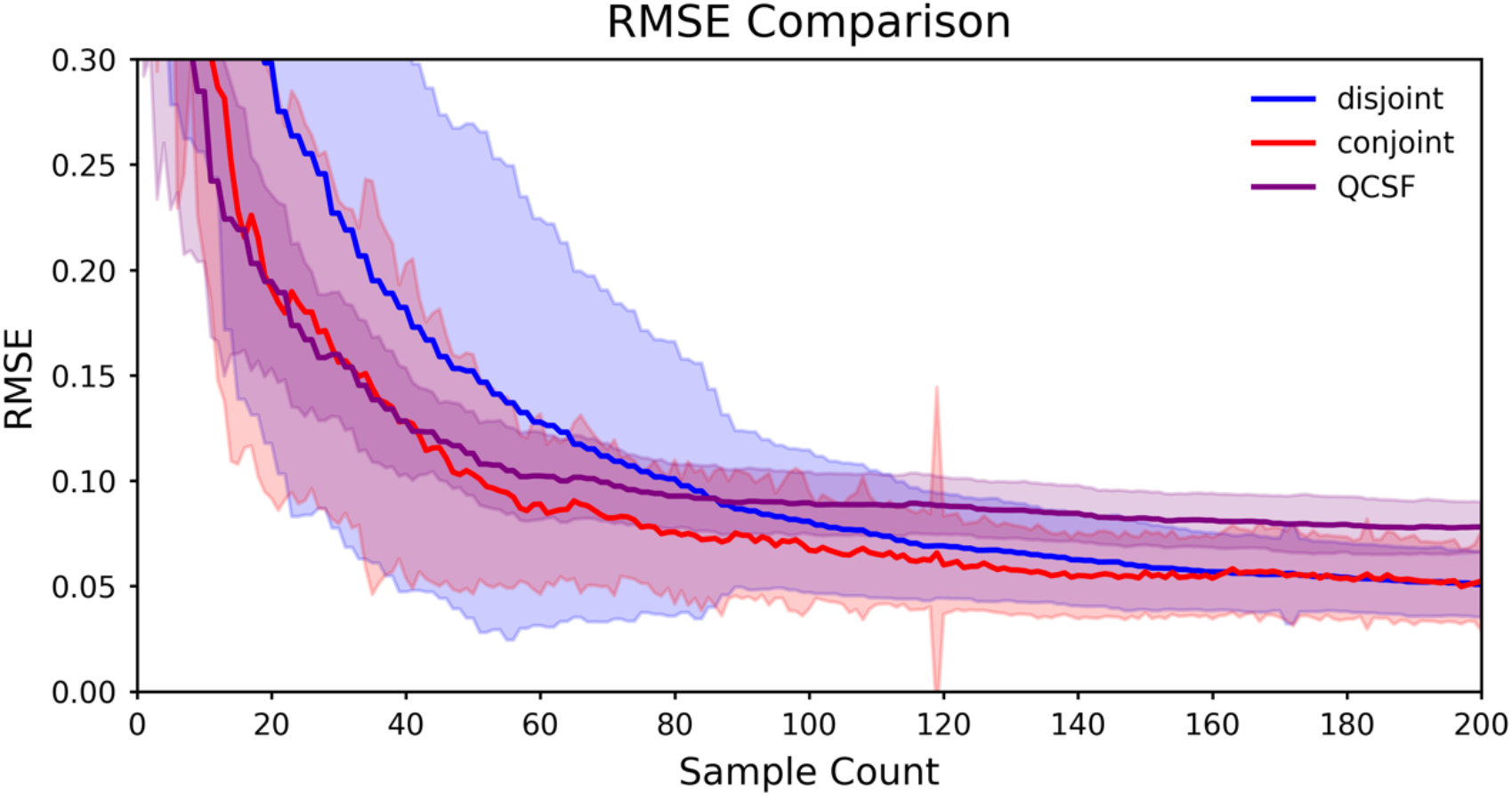
RMSE comparison between second-generation disjoint (blue) and conjoint (red) MLCSF estimates for 210 unique pairs of 20 representative CSF curves acting as ground truth. For comparison, the performance of the quick CSF estimator is also shown (purple). At 119 data points, 1 of the 210 conjoint pairs destabilized its hyperparameters after acquiring a data point, resulting in large average error. The estimate recovered with the immediately following sample. Three of the qCSF phenotype estimates never converged even at 200 samples, and all pairs containing them were removed from analysis.

Numerical quantification of accuracy (i.e., CSF estimate compared to ground truth) and precision (i.e., CSF estimate compared to a repeated estimate) for the two estimators is presented in Table 1. All values are in log contrast units. Estimator precision was estimated directly by repeating estimates of all 210 unique phenotype pairs with a different random seed and quantifying the results. Disjoint and conjoint estimators are both unbiased and accurate, with conjoint estimators achieving higher precision.

**Table 1:** Average differences between CSF estimates and ground truth (accuracy), as well as between repeated CSF estimates (precision) for all 210 unique pairs as a function of active sample count.

| Number of Active Samples | 20 |  | 50 |  | 100 |  |
| --- | --- | --- | --- | --- | --- | --- |
|  | mean | std | mean | std | mean | std |
| **Mean Signed Differences (second – first)** |  |  |  |  |  |  |
| Disjoint – Truth | 0.0273 | 0.3486 | 0.0130 | 0.1915 | –0.0029 | 0.0875 |
| Conjoint – Truth | –0.0027 | 0.2672 | 0.0001 | 0.1419 | –0.0001 | 0.0806 |
| qCSF – Truth | –0.0494 | 0.2210 | 0.0026 | 0.1512 | 0.0099 | 0.1287 |
| Disjoint – Disjoint | 0.0222 | 0.4587 | 0.0048 | 0.2500 | –0.0034 | 0.1174 |
| Conjoint – Conjoint | –0.0108 | 0.3494 | –0.0082 | 0.1862 | 0.0011 | 0.1086 |
| qCSF – qCSF | 0.0035 | 0.3013 | 0.0029 | 0.1972 | 0.0013 | 0.1623 |
| **Absolute Differences** |  |  |  |  |  |  |
| \|Disjoint – Truth\| | 0.2460 | 0.2486 | 0.1195 | 0.1503 | 0.0663 | 0.0572 |
| \|Conjoint – Truth\| | 0.1866 | 0.1913 | 0.0961 | 0.1044 | 0.0623 | 0.0511 |
| \|qCSF – Truth\| | 0.1659 | 0.1542 | 0.1038 | 0.1100 | 0.0829 | 0.0990 |
| \|Disjoint – Disjoint\| | 0.3376 | 0.3113 | 0.1677 | 0.1855 | 0.0898 | 0.0757 |
| \|Conjoint – Conjoint\| | 0.2510 | 0.2433 | 0.1292 | 0.1343 | 0.0828 | 0.0703 |
| \|qCSF – qCSF\| | 0.2218 | 0.2041 | 0.1317 | 0.1468 | 0.0970 | 0.1302 |
| **Root Mean Square Errors/Differences** |  |  |  |  |  |  |
| RMSE(Disjoint, Truth) | 0.3497 |  | 0.1920 |  | 0.0876 |  |
| RMSE(Conjoint, Truth) | 0.2672 |  | 0.1419 |  | 0.0806 |  |
| RMSE(qCSF, Truth) | 0.2265 |  | 0.1512 |  | 0.1291 |  |
| RMSD(Disjoint, Disjoint) | 0.4592 |  | 0.2501 |  | 0.1174 |  |
| RMSD(Conjoint, Conjoint) | 0.3495 |  | 0.1864 |  | 0.1086 |  |
| RMSD(qCSF, qCSF) | 0.3014 |  | 0.1972 |  | 0.1624 |  |

To investigate whether conjoint estimator’s advantage over disjoint estimation decreases for greater phenotypic difference between a pair of CSFs, the analysis of Figure 6 was repeated in subgroups stratified by the cumulate RMSE difference between the pair as sample count increases to 200. This analysis is shown in **Figure 7**. A weak trend (intercept = 3.3×10^−2^; slope = –2.2×10^−3^, 95% CI: [–3.5×10^−3^, –8.0×10^−4^]; *r*^2^ = 0.048) is observable and statistically significant (*t*_208_ = –3.2, *p* = 0.0016, *n* = 210, paired-samples *t* test). Therefore, the efficiency benefit of conjoint estimation is greater with similar ground truth CSFs, but not tremendously so.

**Figure 7:**
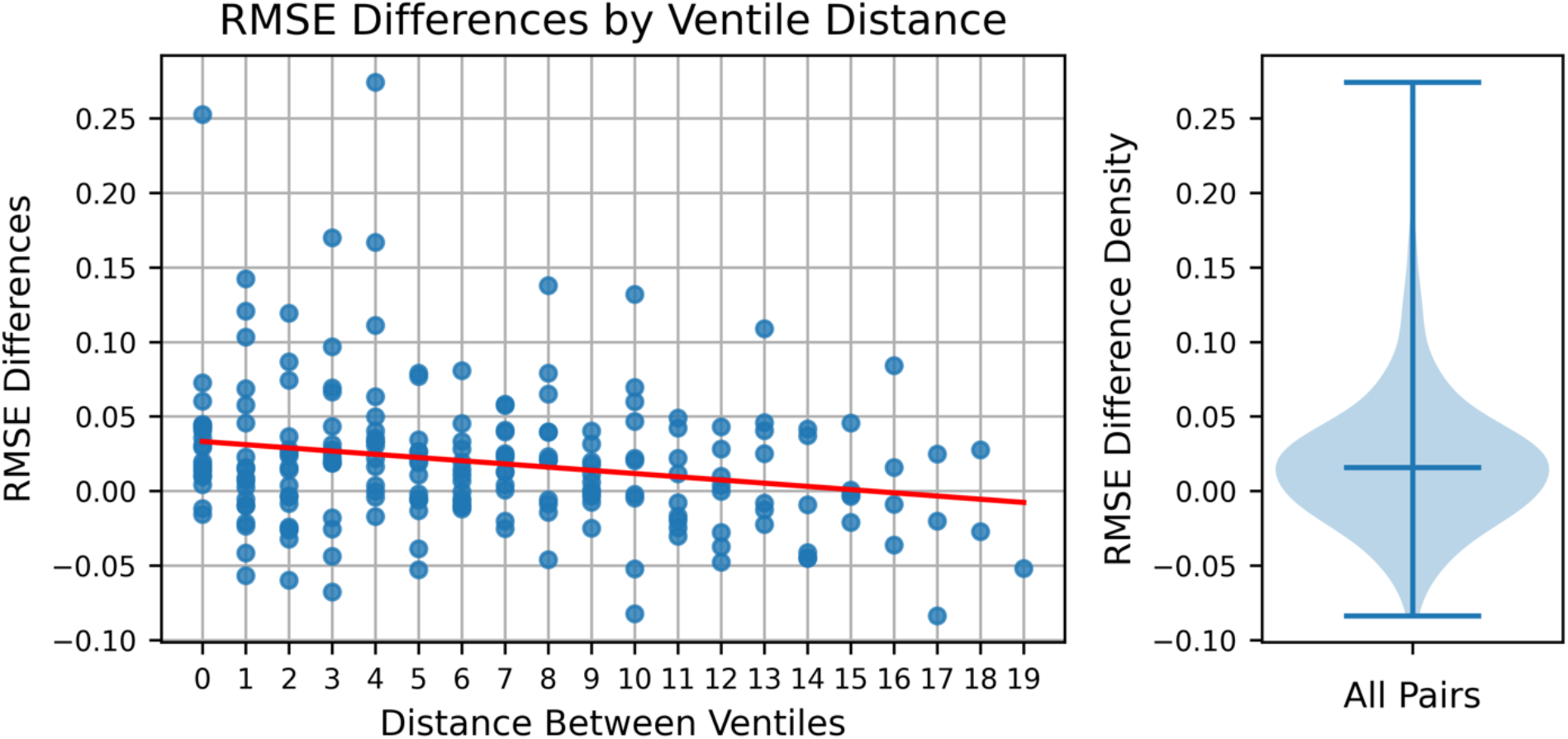
Efficiency gain for second-generation conjoint relative to disjoint MLCSF estimation. RMSE evolution analysis as in Figure 6 was conducted for each of the 210 phenotype pairs. For each pair, the conjoint RMSE evolution curve was subtracted from the disjoint curve up to 100 data samples. The integral of the resulting difference curve formed the RMSE difference between the two estimation methods for that pair. These values were plotted against the distance between the phenotypes in ventiles on the left. The red regression line shows a gradual decrease in the efficiency advantage of conjoint estimation for more dissimilar phenotypes. A violin plot summarizing the entire population distribution is shown on the right with median and range indicated by horizontal lines.

## Discussion

The high degree of flexibility of GPs provides many potential adjustments to improve estimator performance, but also multiple conditions in which such adjustments may be necessary to achieve the full potential of the algorithm. These conditions may differ for different applications. For example, the machine learning audiogram is evaluated, like all audiogram procedures, in relative units. Healthy individuals with no symptoms of hearing loss result in audiogram thresholds near zero for all sound frequencies. Other individuals are compared against that baseline. As a result, even extreme cases of partial peripheral deafferentation result in threshold audiograms of relatively low curvature (Schlittenlacher et al., 2018).

In contrast, CSFs are represented in absolute units and curvatures can be higher at some spatial frequencies and simultaneously lower at other spatial frequencies. The squared exponential kernel in the first-generation MLCSF applies the same curvature constraint at all spatial frequencies. Multiple approaches exist to accommodate this additional knowledge about the nature of the latent function for this application. A typical approach would be to generalize the kernel form to one that provides the additional flexibility of multiple curvatures, including the rational quadratic or Matérn (Rasmussen & Williams, 2006). In fact, the Matérn kernel was selected for machine learning visual field estimation for this reason (Chesley & Barbour, 2020).

While these types of kernel manipulations or even converting absolute CSF units to relative units prior to fitting were considered, a simpler approach became apparent. Because of the reliable concave-down shape of CSFs, adding additional model flexibility to be able to represent closed-path arcs was hypothesized to have utility. The kernel change adopted for second-generation MLCSF would allow circles, ellipses or other closed-form paths to be represented. Such paths are not functions and thus could not represent actual CSFs. Enough flexibility was retained in the second-generation kernel to represent actual CSFs, however, so the empirical question is whether the extra flexibility can improve accuracy without penalizing efficiency. In terms of overall accuracy, Figure 4 reveals that the second-generation kernel performs no worse on average under experiment 1 conditions than the first-generation kernel.

The second-generation model’s systematically improved performance suggests that real-world data may exhibit particularities that disadvantage a strict adherence to strict CSF shapes. The more rigid first-generation model employs a linear kernel in the contrast dimension and a squared exponential kernel in spatial frequency. The more flexible second-generation model, on the other hand, extends the squared exponential kernel to both dimensions. This alteration allows for shared local covariance modeling across both dimensions but relaxes strict CSF shape enforcement as a consequence.

The higher flexibility of the second-generation model compared to the first-generation model is particularly clear at the characteristically steep falloffs at high spatial frequencies. Here, the capabilities of the first-generation model weaken as its assumptions approach invalidity.

Noticeable threshold estimation inaccuracies result, as evidenced in Figure 5. On the other hand, the second-generation model can precisely characterize the entire CSF at both shallower as well as steeper regions. Maintaining accuracy and reliability across a range of conditions underscores the flexible kernel’s superiority in data modeling.

First-generation models also imposed a hard nonlinearity in the acquisition function: the absolute maximum of the utility function. The most obvious consequence of this design is that stimuli tend to cluster in some areas and fail to explore other informative areas simply because the utility function in those areas is slightly smaller. This phenomenon is especially prominent in mid and later stages of active learning when entropy gradients become very shallow with respect to spatial frequency. Thus, imposing the maximum of the utility function routinely leads to quasi-random local distributions of stimuli. Because this issue tended to worsen as data collection continued, the idea to enforce a dispersion of data throughout collection seemed reasonable. This basic alteration successfully distributed stimuli across spatial frequency, leading to coarse shape approximations of ground truth much more quickly.

The conjoint model coregionalization method results in more degrees of freedom than two combined disjoint models. Especially when coupled with the already flexible GP nature, the need to fit additional hyperparameters would conventionally be expected to require more data to achieve good fits, not fewer. Nevertheless, the conjoint MLCSF behavior observed here is consistent with estimator behavior in other domains (Barbour et al., 2018; Heisey et al., 2018).

Early in data collection, the second-generation kernel can produce the unrealistic CSF estimates described earlier. The top panels of **Figure 8** show this phenomenon clearly at 30 samples. Both disjoint CSF estimations in that case curved back upon themselves at high spatial frequencies. With the conjoint estimator, however, this phenomenon does not typically occur and when it does is less dramatic compared to the disjoint estimator. For both estimator types the final estimates are highly accurate.

**Figure 8:**
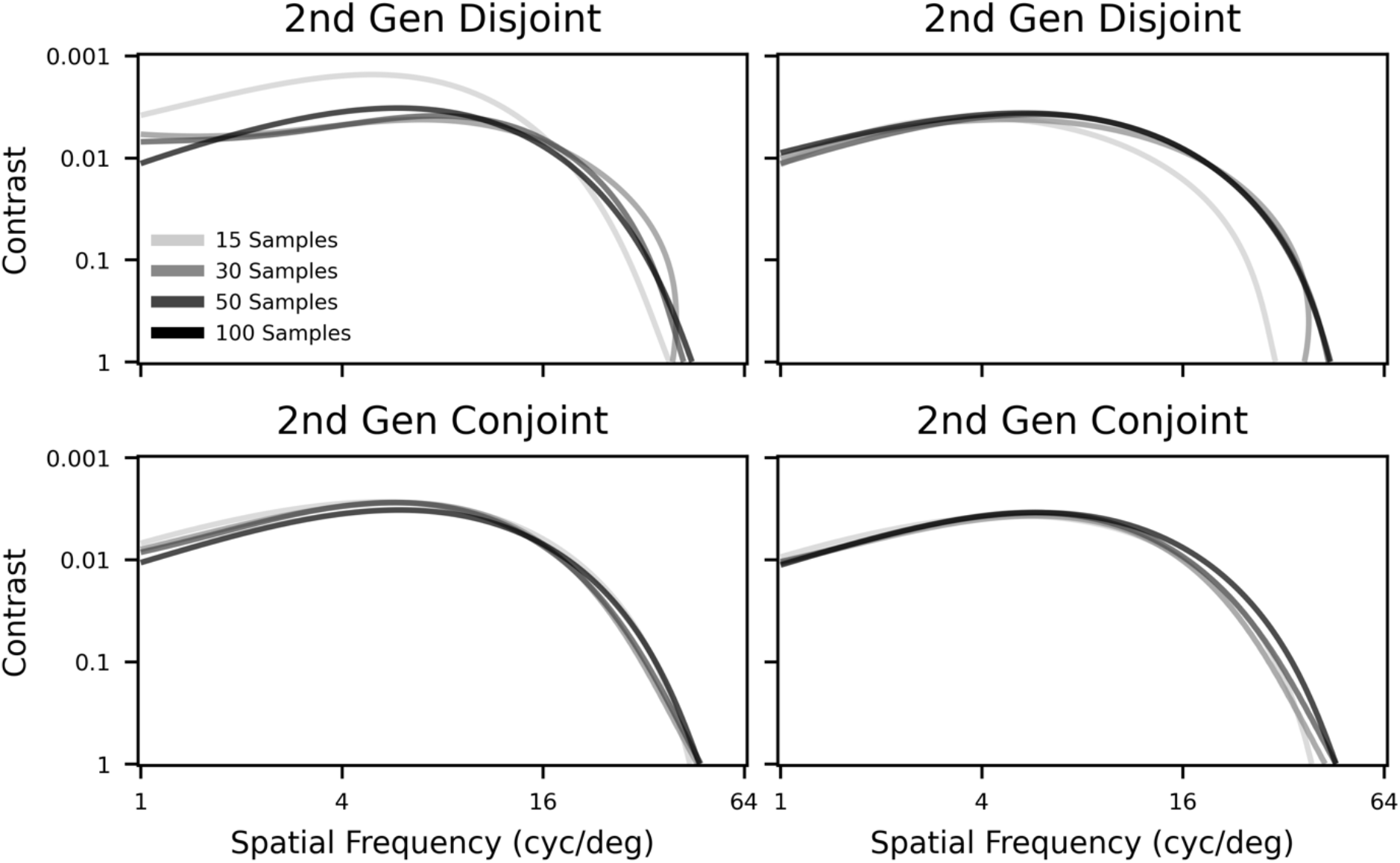
Estimations of second-generation disjoint (top) and conjoint (bottom) MLCSFs estimated at various stages of active learning (15, 30, 50, and 100 samples per task) for a (Normal, Normal) canonical phenotype pair. Conjoint estimation achieves final estimates considerably faster than disjoint estimation. Note that 100 samples per task means 50 samples per phenotype.

Convergence toward final CSF estimate is systematically faster for conjoint estimation compared to disjoint estimation. While this is true on average, Figure 7 shows that there are certainly examples in which disjoint estimation at 100 samples was more accurate than conjoint estimation. Negative RMSE differences indicate when these situations occur. The relative advantage of conjoint estimation steadily declines as the phenotype differences increase. On average, however, conjoint estimation does improve efficiency of CSF estimation.

A slight distinction in clock time also exists between disjoint MLCRF and conjoint MLCRF estimation. Both conditions required steadily increasing model retraining times as data quantity increase, though these values differed for disjoint (e.g., 0.63 ± 0.015 seconds at 10 data points and 1.3 ± 0.036 seconds at 100 data points) and conjoint (e.g., 0.57 ± 0.022 seconds at 10 data points and 1.3 ± 0.038 seconds at 100 data points). Notably, both conditions are twice as fast as the procedure in (Marticorena et al., 2024). Timing computations were made using the CPU on a

Dell Precision 5820 Workstation with Xeon W-2245 8-core 3.90 GHz CPU and 128 GB of RAM.

The collection of improvements between the first and generation MLCRF kernels and acquisition functions reflect the flexibility intrinsic to the GP method. Two modifications—removing the inductive bias of strictly functional CSF shapes and adding a sampling density consideration— systematically sped up convergence rates and improved final estimates across many varying phenotypes. While the present generation of MLCRF may have a high-performing configuration across the included population, the expected CSF form for any particular subpopulation can justify adjustments to the model’s covariance structure or active sampling procedure.

Naive conjoint estimation of two eyes, as demonstrated in this paper, uses the least interesting form of such information: item-level responses from another CSF. In this case, the linear model of coregionalization is free to learn with no priors and no constraints, and thus in large part acts as a mildly informed regularizer. In experimental use with many conditions, however, directed hyperparameter adjustments to the coregionalization matrix could encourage sampling based on expected form shifts, such as the well-known effects of eccentricity or luminance. The flexible coregionalization method can handle non-item-level information such as field of view or visual acuity evaluations to inform CSF estimates and vice versa. This capability opens the door for further developments of semi-parametric, high-speed, low-budget, multi-outcome visual evaluations.

## Conclusions

An updated nonparametric Bayesian estimator learning from simulated contrast behavioral responses has demonstrated improvements in CSF estimation efficiency and accuracy from informed manipulation of model structure and flexibility. These improvements incur no computational penalty when learning two CSFs simultaneously and are scalable to exploit additional data-informed model constraints for further improvements. Future work will evaluate its performance in real-time human data acquisition.

## Data Availability

All data produced in the present study are available upon reasonable request to the authors.

https://github.com/michaeljigo/JigoCarrasco2020

## Acknowledgments

Supported by R21EY033553 and R01EY019693. A patent has been issued covering technology described in this study (Barbour, D. L. et al., 2021).

## Notes

### Competing Interest Statement

The authors have declared no competing interest.

### Funding Statement

This study was funded by R21EY033553.

### Author Declarations

The human data in this manuscript is limited to the openly available data originally located at: https://github.com/michaeljigo/JigoCarrasco2020.

### Summary of Updates

Text has been revised for readability. Additional analysis of the quickCSF estimator was added. Estimator precision was quantified in a new table.

## References

Barbour, D. L., DiLorenzo, J., Sukesan, K. A., Song, X. D., Chen, J. Y., Degen, E. A., Heisey, K. L., & Garnett, R. (2018). Conjoint psychometric field estimation for bilateral audiometry. Behav Res Meth. 10.3758/s13428-018-1062-3

Barbour, D. L., Song, X., Ledbetter, N., Gardner, J., & Weinberger, K. (2021). Fast, Continuous Psychometric Estimation System Utilizing Machine Learning and Associated Method of Use (United States Patent US11037677B2). https://patents.google.com/patent/US11037677B2

Canare, D., Ni, R., & Lu, T. (2019). An open-source implementation of the Quick CSF method. Journal of Vision, 19(10), 86b. 10.1167/19.10.86b

Chesley, B., & Barbour, D. L. (2020). Visual field estimation by probabilistic classification. IEEE Journal of Biomedical and Health Informatics, 24(12), 3499–3506. 10.1109/JBHI.2020.2999567

Chung, S. T. L., & Legge, G. E. (2016). Comparing the shape of contrast sensitivity functions for normal and low vision. Investigative Ophthalmology & Visual Science, 57(1), 198–207. 10.1167/iovs.15-18084

Gardner, J. M., Pleiss, G., Weinberger, K. Q., Bindel, D., & Wilson, A. G. (2018). Gpytorch: Blackbox matrix-matrix gaussian process inference with gpu acceleration. Advances in Neural Information Processing Systems, 31. https://proceedings.neurips.cc/paper_files/paper/2018/file/27e8e17134dd7083b050476733207ea1-Paper.pdf

Goulard, M., & Voltz, M. (1992). Linear coregionalization model: Tools for estimation and choice of cross-variogram matrix. Mathematical Geology, 24(3), 269–286. 10.1007/BF00893750

GPyTorch 1.8.1. (n.d.). Retrieved February 26, 2023, from https://docs.gpytorch.ai/en/stable/

Green, D. M., & Swets, J. A. (1966). Signal Detection Theory and Psychophysics. John Wiley & Sons, Inc.

Gu, H., Kim, W., Hou, F., Lesmes, L. A., Pitt, M. A., Lu, Z.-L., & Myung, J. I. (2016). A hierarchical Bayesian approach to adaptive vision testing: A case study with the contrast sensitivity function. Journal of Vision, 16(6), 15. 10.1167/16.6.15

Heisey, K. L., Buchbinder, J. M., & Barbour, D. L. (2018). Concurrent bilateral audiometric inference. Acta Acustica United with Acustica, 104(5), 762–765. 10.3813/AAA.919218

Hensman, J., Matthews, A., & Ghahramani, Z. (2015). Scalable variational Gaussian process classification. Artificial Intelligence and Statistics, 351–360. http://proceedings.mlr.press/v38/hensman15.pdf

Houlsby, N., Huszár, F., Ghahramani, Z., & Lengyel, M. (2011). Bayesian active learning for classification and preference learning. arXiv Preprint arXiv:1112.5745. 10.48550/arXiv.1112.5745

Jigo, M., & Carrasco, M. (2020). Differential impact of exogenous and endogenous attention on the contrast sensitivity function across eccentricity. Journal of Vision, 20(6), 11. 10.1167/jov.20.6.11

Kalloniatis, M., & Luu, C. (1995). Visual Acuity. In H. Kolb, E. Fernandez, & R. Nelson (Eds.), Webvision: The Organization of the Retina and Visual System. University of Utah Health Sciences Center. http://www.ncbi.nlm.nih.gov/books/NBK11509/

Lesmes, L. A., Lu, Z.-L., Baek, J., & Albright, T. D. (2010). Bayesian adaptive estimation of the contrast sensitivity function: The quick CSF method. Journal of Vision, 10(3), 17. 1-21. 10.1167/10.3.17

Marticorena, D. C. P., Wong, Q. W., Browning, J., Wilbur, K., Jayakumar, S., Davey, P. G., Seitz, A. R., Gardner, J. R., & Barbour, D. L. (2024). Contrast response function estimation with nonparametric Bayesian active learning. Journal of Vision, 24(1), 6. 10.1167/jov.24.1.6

Python 3.10.9. (n.d.). Retrieved January 12, 2024, from https://docs.python.org/3.10/

PyTorch 1.13.1. (n.d.). Retrieved February 26, 2023, from https://pytorch.org/docs/stable/index.html

Rasmussen, C. E., & Williams, C. K. I. (2006). Gaussian Processes for Machine Learning. The MIT Press.

Rohaly, A. M., & Owsley, C. (1993). Modeling the contrast-sensitivity functions of older adults. JOSA A, 10(7), 1591–1599. 10.1364/JOSAA.10.001591

Schlittenlacher, J., Turner, R. E., & Moore, B. C. (2018). A hearing-model-based active-learning test for the determination of dead regions. Trends in Hearing, 22, 2331216518788215. 10.1177/2331216518788215

Song, X. D., Sukesan, K. A., & Barbour, D. L. (2018). Bayesian active probabilistic classification for psychometric field estimation. Attention, Perception & Psychophysics, 80(3), 798–812. 10.3758/s13414-017-1460-0

Song, X. D., Wallace, B. M., Gardner, J. R., Ledbetter, N. M., Weinberger, K. Q., & Barbour, D. L. (2015). Fast, continuous audiogram estimation using machine learning. Ear and Hearing, 36(6), e326–335. 10.1097/AUD.0000000000000186

Tahir, H. J., Parry, N. R. A., Pallikaris, A., & Murray, I. J. (2009). Higher-order aberrations produce orientation-specific notches in the defocused contrast sensitivity function. Journal of Vision, 9(7), 11. 10.1167/9.7.11

Titsias, M. (2009). Variational learning of inducing variables in sparse Gaussian processes. Artificial Intelligence and Statistics, 567–574. https://proceedings.mlr.press/v5/titsias09a.html

Wang, X., Wang, H., Huang, J., Zhou, Y., & Tzvetanov, T. (2016). Bayesian Inference of Two-Dimensional Contrast Sensitivity Function from Data Obtained with Classical One-Dimensional Algorithms Is Efficient. Frontiers in Neuroscience, 10, 616. 10.3389/fnins.2016.00616

Wichmann, F. A., & Hill, N. J. (2001). The psychometric function: I. Fitting, sampling, and goodness of fit. Perception & Psychophysics, 63(8), 1293–1313. 10.3758/BF03194544

Woods, R. L., Bradley, A., & Atchison, D. A. (1996). Consequences of monocular diplopia for the contrast sensitivity function. Vision Research, 36(22), 3587–3596. 10.1016/0042-6989(96)00091-0

Zhao, Y., Lesmes, L. A., Hou, F., & Lu, Z.-L. (2021). Hierarchical Bayesian modeling of contrast sensitivity functions in a within-subject design. Journal of Vision, 21(12), 9. 10.1167/jov.21.12.9

